# Data Driven Approach to Characterize Rapid Decline in Autosomal Dominant Polycystic Kidney Disease

**DOI:** 10.1101/2024.01.26.24301848

**Authors:** John J. Sim, Yu-Hsiang Shu, Simran K. Bhandari, Qiaoling Chen, Teresa N. Harrison, Min Young Lee, Mercedes A. Munis, Kerresa Morrissette, Shirin Sundar, Kristin Pareja, Ali Nourbakhsh, Cynthia J. Willey

## Abstract

**Background:** Autosomal dominant polycystic kidney disease (ADPKD) is a genetic kidney disease with high phenotypic variability. Insights into ADPKD progression could lead to earlier detection and management prior to end stage kidney disease (ESKD). We sought to identify patients with rapid decline (RD) in kidney function and to determine clinical factors associated with RD using a data-driven approach.

**Methods:** A retrospective cohort study was performed among patients with incident ADPKD (1/1/2002-12/31/2018). Latent class mixed models were used to identify RD patients using rapidly declining eGFR trajectories over time. Predictors of RD were selected based on agreements among feature selection methods, including logistic, regularized, and random forest modeling. The final model was built on the selected predictors and clinically relevant covariates.

**Results:** Among 1,744 patients with incident ADPKD, 125 (7%) were identified as RD. Feature selection included 42 clinical measurements for adaptation with multiple imputations; mean (SD) eGFR was 85.2 (47.3) and 72.9 (34.4) in the RD and non-RD groups, respectively. Multiple imputed datasets identified variables as important features to distinguish RD and non-RD groups with the final prediction model determined as a balance between area under the curve (AUC) and clinical relevance which included 6 predictors: age, sex, hypertension, cerebrovascular disease, hemoglobin, and proteinuria. Results showed 72%-sensitivity, 70%-specificity, 70%-accuracy, and 0.77-AUC in identifying RD. 5-year ESKD rates were 38% and 7% among RD and non-RD groups, respectively.

**Conclusion:** Using real-world routine clinical data among patients with incident ADPKD, we observed that six variables highly predicted RD in kidney function.

## Background

Autosomal dominant polycystic kidney disease (ADPKD) is a leading cause of genetic kidney disease with an estimated prevalence of 30-50 per 100,000 persons.^1–4^ It is the fourth leading cause of end stage kidney disease (ESKD) accounting for 5% of ESKD in the United States.^2, 5, 6^ The natural progression of the disease is characterized by pathogenic genetic mutations that lead to fluid filled cysts resulting in irreversible damage to kidney parenchyma and loss of kidney function.^7, 8^ ADPKD is genetically heterogenous and is caused by pathogenic mutations most commonly in the PKD1 (85% of cases) and PKD2 (15% of cases) genes. There is interfamilial and intrafamilial variability in the natural course of ADPKD.^9–13^ While 50% of patients progress to ESKD by their sixth decade of life, the decline in eGFR typically occurs rapidly later in adulthood due to initial compensatory glomerular hyperfiltration. While eGFR is an established marker of kidney function, it alone has not shown to reliably assess ADPKD burden nor prognosticate outcomes.^14–16^

Current methods to identify rapid progression in ADPKD have relied on resource intensive prognostic approaches and tools. One of the first methods was developed using data from the Consortium for Radiologic Imaging Studies of Polycystic Kidney Disease (CRISP) cohort and Mayo database. Mayo Imaging Classification (MIC) necessitates imaging to determine height adjusted total kidney volume (htTKV), along with age, race, and sex to predict time to decline in eGFR.^7^ While race is used to calculate eGFR, it is not factored into determining risk of progression to ESKD. The Predicting Renal Outcome in PKD (PROPKD) score uses genotype, sex, and the presence of a urologic event and/or hypertension before the age of 35 to predict onset of ESKD.^17, 18^ The PROPKD score relies on genotype results limiting its utility. Both the MIC and the PROPKD score were derived from primarily Caucasian populations and thus may be limited in generalizability.^8, 19^ We previously characterized and described an ADPKD cohort and observed racial/ethnic differences in the proportion of patients with kidney failure, age of kidney failure onset, and likelihood of having had kidney transplantation.^20^ Thus, approaches to identify rapid progressors early in the disease course with readily available clinical data and considering population diversity are warranted.

Using a large diverse population of patients with ADPKD and longitudinal eGFR data collected within a routine clinical care environment, we sought to identify rapid decline (RD) in kidney function and determine clinical factors associated with RD using a data-driven approach.

## Material and Methods

### Study Population

A retrospective cohort study of Kaiser Permanente Southern California (KPSC) members between January 1, 2002 to December 31, 2018 was performed. KPSC is an integrated health system comprised of 15 medical centers and over 230 satellite clinics providing care to over 4.8 million members. The membership population is racially, ethnically, and socioeconomically diverse, reflecting the general population of Southern California.^21^ Complete healthcare encounters are tracked using a comprehensive electronic health record (EHR) from which all study information were extracted.

The study population from which this cohort was identified has been previously described.^4, 18^ In brief, the study population included patients of any age with a minimum of 1-year continuous membership in the health plan. This time requirement was used to reliably capture incident ADPKD diagnoses and comorbidities. Inpatient and outpatient *International Classifications of Diseases, Ninth and Tenth Revision (ICD-9, ICD-10)* ADPKD diagnoses codes (ICD-9: 753.12, 753.13; ICD-10: Q61.2, Q61.3) were used to identify patients.

Incident ADPKD was defined as newly diagnosed and not having a prior diagnosis of ADPKD. Patients were required to have ≥ 2 diagnosis codes on 2 separate encounter dates (which may have been consecutive days) from inpatient, emergency department (ED), or ambulatory care settings. Patients were excluded if they had a prior diagnosis of ADPKD, ≥ 2 diagnosis codes for autosomal recessive polycystic kidney disease (ARPKD) or did not have 1-year of continuous KPSC membership. Patients with ESKD (defined as treatment with dialysis or kidney transplant), eGFR <15mL/min/1.73m^2^, or no eGFR information at baseline and during follow-up were also excluded (Supplementary Figure 1). Patients were followed until they experienced ESKD, death, disenrollment from the healthcare plan, or until the end of the observation period (January 31, 2020).

### Data Collection

Information on demographics, clinical characteristics, and medications were obtained for patients with incident ADPKD in the 1 year prior to the index date. All laboratory data, vital sign assessments (including blood pressure measurements and body mass index), and diagnostic and procedure codes were extracted from the EHR. Kidney function was expressed as eGFR calculated from serum creatinine levels using the 2009 Chronic Kidney Disease Epidemiology Collaboration equation.^22^ Proteinuria was defined as urinalysis positive for protein, urine protein/creatinine ratio >0.2 g/g, urine albumin/creatinine ratio >30mg/g, or a 24-hour urine collection with >200mg total protein or >30mg of albumin. ESKD was defined as treatment with hemodialysis, peritoneal dialysis, or kidney transplant.

Medication use was retrieved from internal pharmacy dispensing records. Health care utilization and an ever history of comorbidities (diabetes mellitus (DM), hypertension (HTN), hyperlipidemia, ischemic heart disease (ICH), congestive heart failure (CHF), cerebrovascular disease, urologic diseases, abdominal pain, and liver disease) were extracted from the EHR. The Elixhauser Comorbidity Index was also extracted from the EHR categorizing 31 comorbidities using diagnosis codes with each comorbidity being assigned a value of ‘1’.^23^ Data on hospitalizations and diagnoses that occurred outside of the KPSC healthcare system were available through administrative claims records. KPSC death records were obtained by identifying death that occurred within KPSC-owned facilities.^24^

### Analysis

Descriptive statistics stratified by the RD vs. non-rapid decliner groups were used to report demographic and clinical characteristics among patients with ADPKD. The standardized mean difference (SMD) was used to test for distance between group means (i.e., effect size). An SMD > 0.1 indicates the distributions between the two groups are unbalanced.

Serial eGFRs were evaluated for each patient to determine the pattern of eGFR change over time. Group-based trajectory analysis was applied to identify possible patterns in potential distinct trajectory groups based on the longitudinal eGFR data using a latent class mixed model (LCMM). LCMM was performed by regressing eGFR on the measurement time assuming random effects of time between individuals. The appropriate assumptions of trajectories were explored with linear, beta, and spline distributions. For each distribution assumption, the number of distinct trajectory groups were evaluated from 2 to 5 in consideration of the sufficient sample size in each group. The best fitted results were determined based on the smallest model Akaike Information Criterion (AIC). After distinct trajectory groups were identified in the analysis, the trajectory curves of each group were visualized using a locally weighted scatterplot smoothing (LOWESS) approach. Patients with ADPKD showing the steepest decline in kidney function were categorized in one (or more) trajectory curve(s) and were defined as the RD group. Accordingly, the other group(s) was defined as the non-rapid decliner (non-RD).

After RD patients were identified based on the longitudinal eGFR data using LCMM, the next steps explored potential associations included baseline demographics, vital signs, comorbidities, and laboratory data. Given the numerous baseline variables that were considered as potential predictors for RD, several feature selection steps were performed to help identify the variables with higher importance. Variables with missingness above 30% were initially included, and the missing data were addressed using multiple imputation by chained equations (MICE). To achieve better reliability, three feature selection methods were applied: logistic regression, Least Absolute Shrinkage and Selection Operator (LASSO) regression, and random forest. The important features were determined based on mutual agreements between the three methods. In the logistic model, variables were selected if the p-values were less than 0.05 using likelihood ratio test. In LASSO, lambda parameter was tuned to find the top 80th percentile of important features as the variables with non-zero coefficients. In random forest, variables at top 80th percentile importance were selected. After performing the three methods in ten sets of the imputed data, pooled statistics were calculated to determine the selected variables in each method (pooled p-values in logistic model, grouped adaptive LASSO, and averaged-scaled importance in random forest). Finally, variables with mutual agreements of being selected between the methods were included as the predictors in building the prediction model based on the non-missing data.

The logistic model was used to build the prediction model for RD due to its accessibility and simplicity. Variables selected from the feature selection were included in the model, and the prediction performance was evaluated using area under the curve (AUC). A few additional clinical variables of interest were included in the model to see if the prediction performance could be improved, even if they were not initially selected in the feature selection steps. The final prediction model was determined based on the balance between model simplicity, AUC, and clinical relevance.

Time to ESKD and/or mortality were the primary outcomes. ESKD rates overall and before age 53 (50^th^ percentile), 60 (based in the PROPKD score), 62 (based on Mayo Clinic Research), and 63 (75^th^ percentile) were evaluated. ESKD rates within 5 years of incident ADPKD were evaluated for risk assessment among incident ADPKD patients.

Descriptive statistics and cumulative incidence plots were used to describe time to mortality and ESKD stratified by RD status for each of the outcomes of interest. The incidence rate (IR) per 1,000 person-years and the 95% confidence intervals (CI) were estimated using Poisson regression with robust standard error.

All statistical analyses were generated using the SAS Enterprise Guide (version 9.4; SAS Institute, Cary, North Carolina, USA). The KPSC Institutional Review Board (IRB) reviewed and approved the protocol of this study (#11823). A waiver of informed consent was obtained due to the retrospective nature of this study. Data were accessed for research purposes for this study during the period 01/06/2019 through 01/12/2022.

## Results

### Study Population

A total of 1,744 patients with incident ADPKD were included in the study. The mean (SD) age of the study population was 50.6 (19.1) years, 52.4% males, 41.6% White, 13.8% Black, 31.5% Hispanic, and 9.9% Asian/Pacific Islander (Table 1). Mean blood pressure was 129/76 mm Hg and mean (SD) eGFR was 73.8 (35.6) mL/min/1.73m^2^. At baseline, 60.0% of patients had a history of HTN, 33.0% had a history of urologic disease, and 12.6% had a history of DM.

**Table 1.**
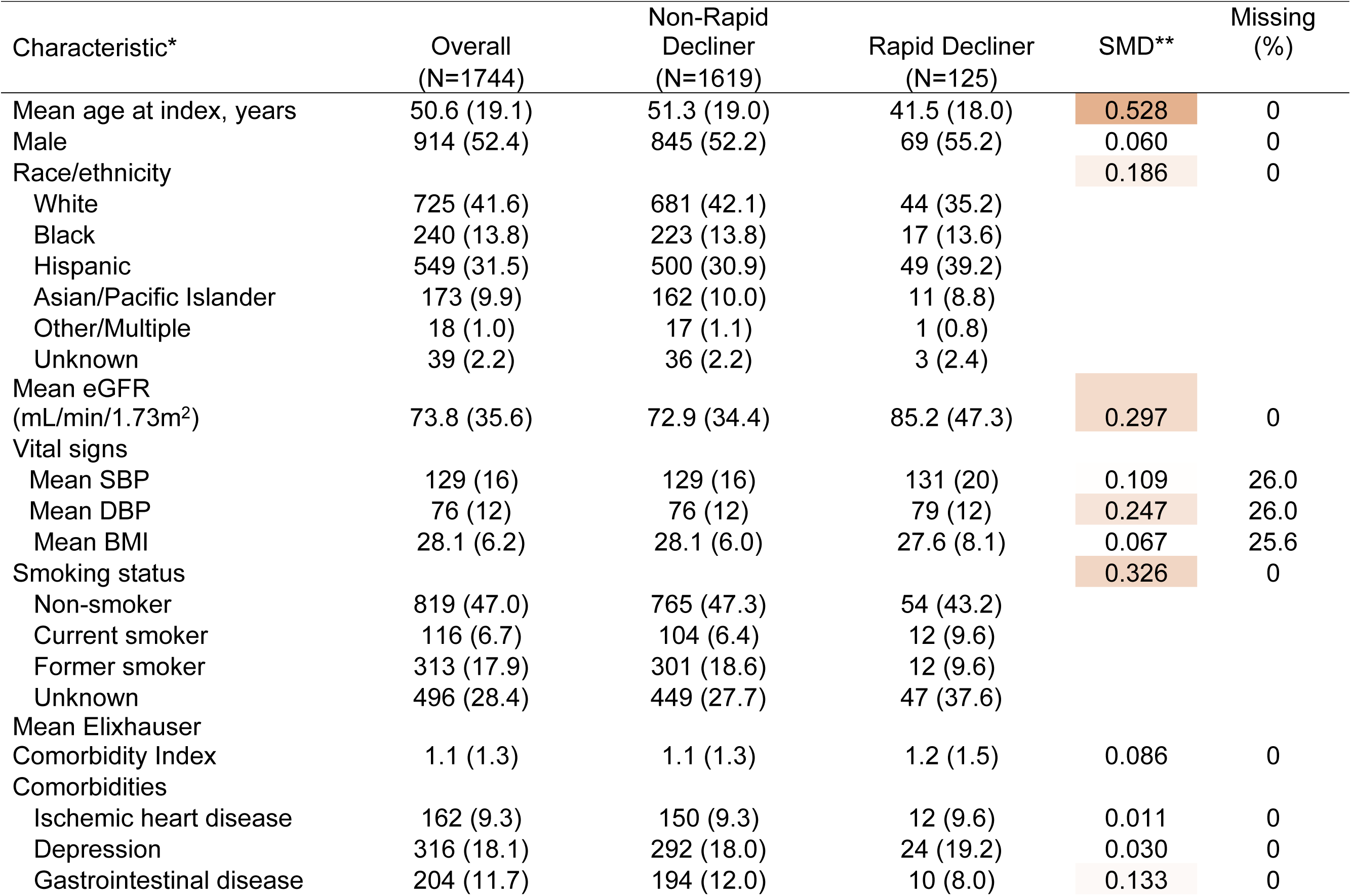
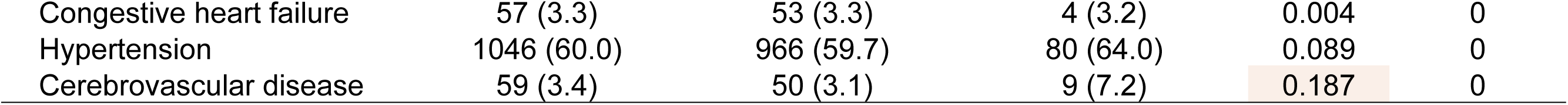
Baseline Characteristics and Comorbidities Among Patients in the Non-Rapid Decliner vs. Rapid Decliner Group.

| Characteristic* | Overall<br>(N=1744) | Non-Rapid<br>Decliner<br>(N=1619) | Rapid Decliner<br>(N=125) | SMD** | Missing<br>(%) |
| --- | --- | --- | --- | --- | --- |
| Mean age at index, years | 50.6 (19.1) | 51.3 (19.0) | 41.5 (18.0) | 0.528 | 0 |
| Male | 914 (52.4) | 845 (52.2) | 69 (55.2) | 0.060 | 0 |
| Race/ethnicity |  |  |  | 0.186 | 0 |
| White | 725 (41.6) | 681 (42.1) | 44 (35.2) |  |  |
| Black | 240 (13.8) | 223 (13.8) | 17 (13.6) |  |  |
| Hispanic | 549 (31.5) | 500 (30.9) | 49 (39.2) |  |  |
| Asian/Pacific Islander | 173 (9.9) | 162 (10.0) | 11 (8.8) |  |  |
| Other/Multiple | 18 (1.0) | 17 (1.1) | 1 (0.8) |  |  |
| Unknown | 39 (2.2) | 36 (2.2) | 3 (2.4) |  |  |
| Mean eGFR<br>(mL/min/1.73m<sup>2</sup>) | 73.8 (35.6) | 72.9 (34.4) | 85.2 (47.3) | 0.297 | 0 |
| Vital signs |  |  |  |  |  |
| Mean SBP | 129 (16) | 129 (16) | 131 (20) | 0.109 | 26.0 |
| Mean DBP | 76 (12) | 76 (12) | 79 (12) | 0.247 | 26.0 |
| Mean BMI | 28.1 (6.2) | 28.1 (6.0) | 27.6 (8.1) | 0.067 | 25.6 |
| Smoking status |  |  |  | 0.326 | 0 |
| Non-smoker | 819 (47.0) | 765 (47.3) | 54 (43.2) |  |  |
| Current smoker | 116 (6.7) | 104 (6.4) | 12 (9.6) |  |  |
| Former smoker | 313 (17.9) | 301 (18.6) | 12 (9.6) |  |  |
| Unknown | 496 (28.4) | 449 (27.7) | 47 (37.6) |  |  |
| Mean Elixhauser<br>Comorbidity Index | 1.1 (1.3) | 1.1 (1.3) | 1.2 (1.5) | 0.086 | 0 |
| Comorbidities |  |  |  |  |  |
| Ischemic heart disease | 162 (9.3) | 150 (9.3) | 12 (9.6) | 0.011 | 0 |
| Depression | 316 (18.1) | 292 (18.0) | 24 (19.2) | 0.030 | 0 |
| Gastrointestinal disease | 204 (11.7) | 194 (12.0) | 10 (8.0) | 0.133 | 0 |
| Congestive heart failure | 57 (3.3) | 53 (3.3) | 4 (3.2) | 0.004 | 0 |
| Hypertension | 1046 (60.0) | 966 (59.7) | 80 (64.0) | 0.089 | 0 |
| Cerebrovascular disease | 59 (3.4) | 50 (3.1) | 9 (7.2) | 0.187 | 0 |

### RD vs non-RD

Based on the best AIC model, we identified two groups in the longitudinal eGFR data: 125 (7%) patients in the RD group and 1,619 (93%) patients in the non-RD group (Figure 1). Patients in the RD group had a mean (SD) age of 41.5 (18.0) years vs. 51.3 (19.0) years in the non-RD group. Compared to non-RD, the RD group had a higher proportion of patients who were male (55.2% vs 52.2%), Hispanic (39.2% vs 30.9%), had higher baseline BP (131/79 mm Hg vs 129/76 mm Hg), and were more likely to have HTN (64.0% vs 59.7%) and cerebrovascular disease (7.2% vs 3.1%) (Table 1).

**Figure 1:** Trajectory groups using latent class mixed model assuming spline distribution.

History of urologic disease and abdominal pain were lower in the RD group (28.0% vs. 33.4%, 40.8% vs. 51.1%, respectively). Mean (SD) baseline eGFR was 85.2 (47.3) and 72.9 (34.4) mL/min/1.73m^2^ in the RD and non-RD groups, respectively. Baseline sodium, bicarbonate, BUN, vitamin D, HDL, CRP, PTH and albumin were lower in RD compared to non-RD (Supplemental Table 1). Baseline hemoglobin was 13.1 (1.6) g/dL and 13.6 (1.7) g/dL the in RD and non-RD groups, respectively. The RD group was found to have higher urinary protein, lower urinary phosphorus, and lower urinary calcium compared to non-RD group.

### Predictors of Rapid Decline

Of the 42 variables assessed, 7 were found to have significant agreement in identifying RD including: age, hemoglobin, creatinine, proteinuria, hypertension, cerebrovascular disease, and liver disease (Table 2). In addition, 4 pre-selected clinically relevant variables were identified and included in the model: sex, race/ethnicity, diabetes, and history of urologic events. Based on a balance between clinical relevance and AUC assessed using non-missing data, creatinine and history of liver disease were excluded and sex was included in the final model. The 6 final predictors of RD were younger age at onset [Odds Ratio (OR) 0.941 (95% CI, 0.927, 0.955)], male sex [OR 1.795 (95% CI, 1.129, 2.879)], hypertension [OR 4.402 (95% CI, 2.469, 8.059)], cerebrovascular disease [OR 3.612 (95% CI, 1.378, 8.399)], a 1g/dL increase of hemoglobin [OR 0.820 (95% CI, 0.719, 0.936)], and proteinuria [OR 2.887 (95% CI, 1.861, 4.536)] (Table 3). Results showed 72% sensitivity, 70% specificity, 70% accuracy, and 0.77 AUC in identifying the RD group. AUC results were considered fair for values between 0.7 and 0.8 (Figure 2).

**Figure 2:** Area under the receiver operating characteristic curve.

**Table 2.**
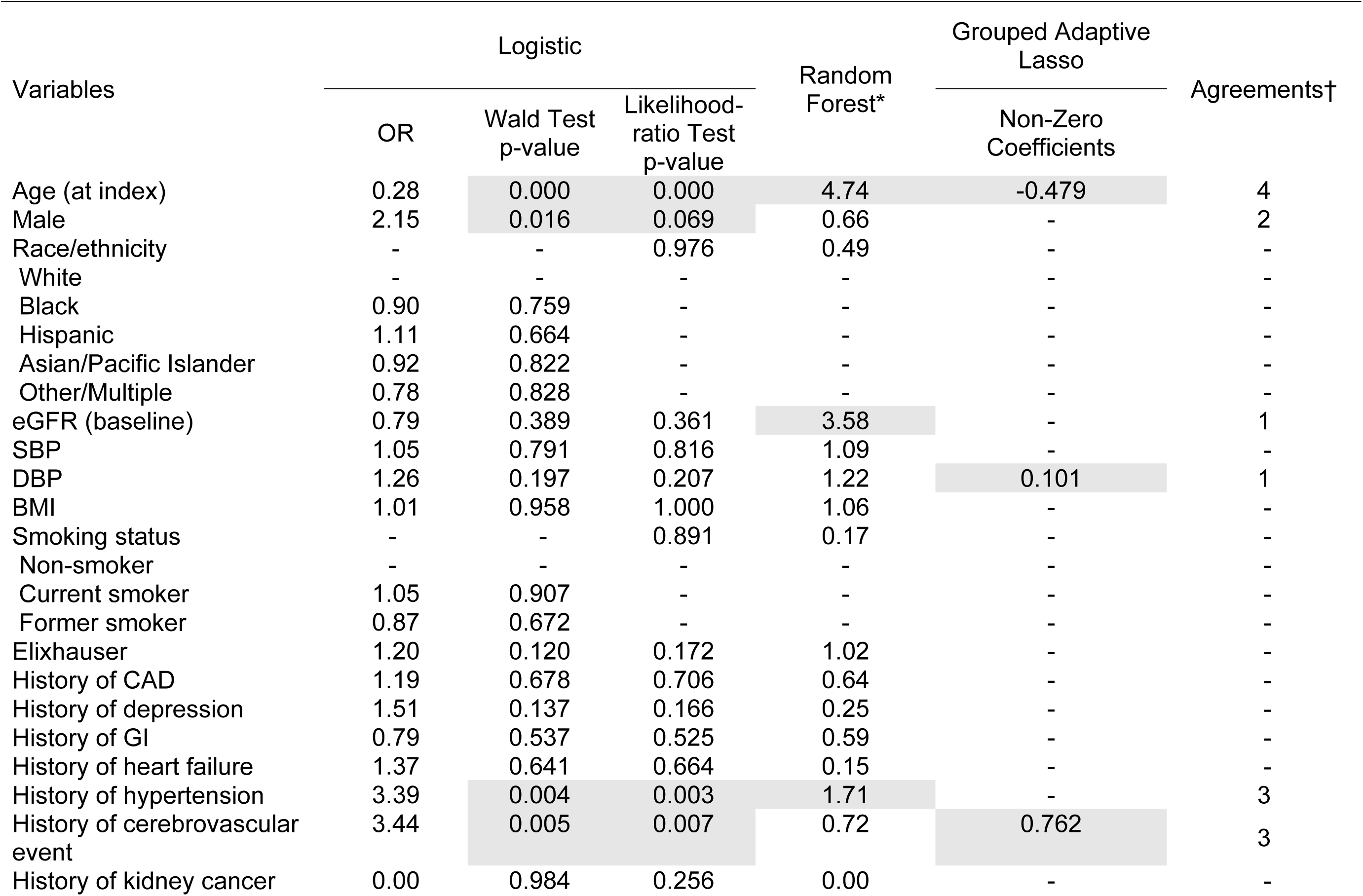
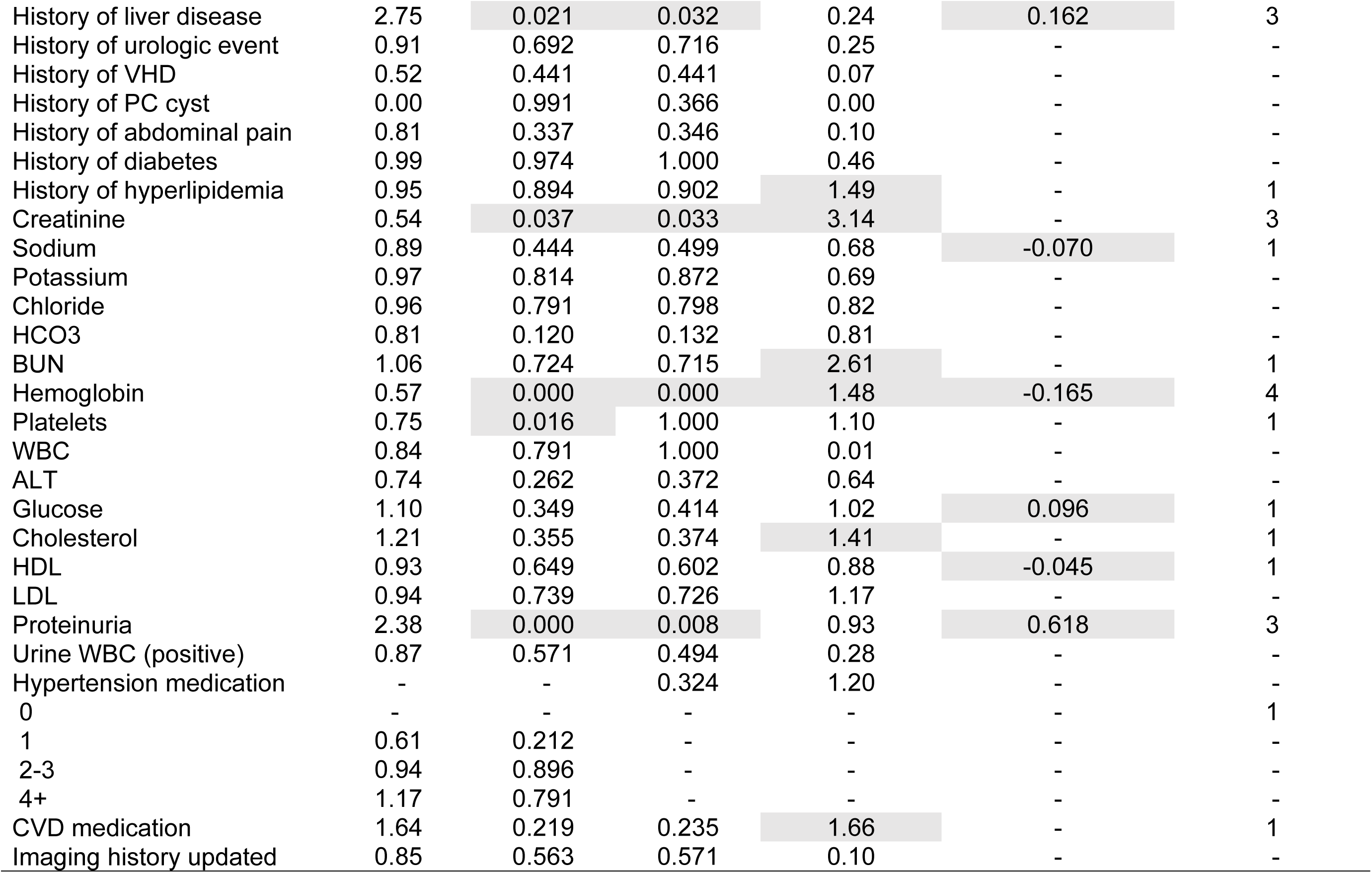
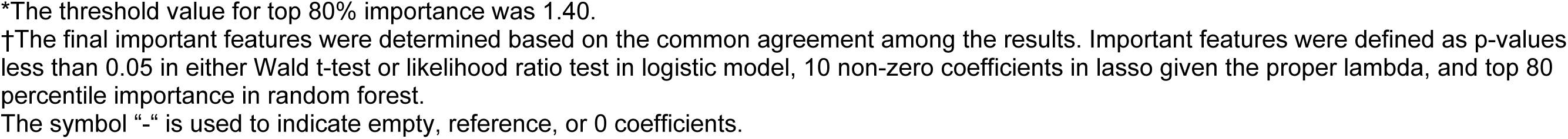
Results of Feature Selection Approaches in Multiple Imputed Datasets.

| Variables | Logistic |  |  | Random Forest* | Grouped Adaptive Lasso | Agreements† |
| --- | --- | --- | --- | --- | --- | --- |
|  | OR | Wald Test p-value | Likelihood-ratio Test p-value |  | Non-Zero Coefficients |  |
| Age (at index) | 0.28 | 0.000 | 0.000 | 4.74 | -0.479 | 4 |
| Male | 2.15 | 0.016 | 0.069 | 0.66 | - | 2 |
| Race/ethnicity | - | - | 0.976 | 0.49 | - | - |
| White | - | - | - | - | - | - |
| Black | 0.90 | 0.759 | - | - | - | - |
| Hispanic | 1.11 | 0.664 | - | - | - | - |
| Asian/Pacific Islander | 0.92 | 0.822 | - | - | - | - |
| Other/Multiple | 0.78 | 0.828 | - | - | - | - |
| eGFR (baseline) | 0.79 | 0.389 | 0.361 | 3.58 | - | 1 |
| SBP | 1.05 | 0.791 | 0.816 | 1.09 | - | - |
| DBP | 1.26 | 0.197 | 0.207 | 1.22 | 0.101 | 1 |
| BMI | 1.01 | 0.958 | 1.000 | 1.06 | - | - |
| Smoking status | - | - | 0.891 | 0.17 | - | - |
| Non-smoker | - | - | - | - | - | - |
| Current smoker | 1.05 | 0.907 | - | - | - | - |
| Former smoker | 0.87 | 0.672 | - | - | - | - |
| Elixhauser | 1.20 | 0.120 | 0.172 | 1.02 | - | - |
| History of CAD | 1.19 | 0.678 | 0.706 | 0.64 | - | - |
| History of depression | 1.51 | 0.137 | 0.166 | 0.25 | - | - |
| History of GI | 0.79 | 0.537 | 0.525 | 0.59 | - | - |
| History of heart failure | 1.37 | 0.641 | 0.664 | 0.15 | - | - |
| History of hypertension | 3.39 | 0.004 | 0.003 | 1.71 | - | 3 |
| History of cerebrovascular event | 3.44 | 0.005 | 0.007 | 0.72 | 0.762 | 3 |
| History of kidney cancer | 0.00 | 0.984 | 0.256 | 0.00 | - | - |
| History of liver disease | 2.75 | 0.021 | 0.032 | 0.24 | 0.162 | 3 |
| History of urologic event | 0.91 | 0.692 | 0.716 | 0.25 | - | - |
| History of VHD | 0.52 | 0.441 | 0.441 | 0.07 | - | - |
| History of PC cyst | 0.00 | 0.991 | 0.366 | 0.00 | - | - |
| History of abdominal pain | 0.81 | 0.337 | 0.346 | 0.10 | - | - |
| History of diabetes | 0.99 | 0.974 | 1.000 | 0.46 | - | - |
| History of hyperlipidemia | 0.95 | 0.894 | 0.902 | 1.49 | - | 1 |
| Creatinine | 0.54 | 0.037 | 0.033 | 3.14 | - | 3 |
| Sodium | 0.89 | 0.444 | 0.499 | 0.68 | -0.070 | 1 |
| Potassium | 0.97 | 0.814 | 0.872 | 0.69 | - | - |
| Chloride | 0.96 | 0.791 | 0.798 | 0.82 | - | - |
| HCO3 | 0.81 | 0.120 | 0.132 | 0.81 | - | - |
| BUN | 1.06 | 0.724 | 0.715 | 2.61 | - | 1 |
| Hemoglobin | 0.57 | 0.000 | 0.000 | 1.48 | -0.165 | 4 |
| Platelets | 0.75 | 0.016 | 1.000 | 1.10 | - | 1 |
| WBC | 0.84 | 0.791 | 1.000 | 0.01 | - | - |
| ALT | 0.74 | 0.262 | 0.372 | 0.64 | - | - |
| Glucose | 1.10 | 0.349 | 0.414 | 1.02 | 0.096 | 1 |
| Cholesterol | 1.21 | 0.355 | 0.374 | 1.41 | - | 1 |
| HDL | 0.93 | 0.649 | 0.602 | 0.88 | -0.045 | 1 |
| LDL | 0.94 | 0.739 | 0.726 | 1.17 | - | - |
| Proteinuria | 2.38 | 0.000 | 0.008 | 0.93 | 0.618 | 3 |
| Urine WBC (positive) | 0.87 | 0.571 | 0.494 | 0.28 | - | - |
| Hypertension medication | - | - | 0.324 | 1.20 | - | - |
| 0 | - | - | - | - | - | 1 |
| 1 | 0.61 | 0.212 | - | - | - | - |
| 2-3 | 0.94 | 0.896 | - | - | - | - |
| 4+ | 1.17 | 0.791 | - | - | - | - |
| CVD medication | 1.64 | 0.219 | 0.235 | 1.66 | - | 1 |
| Imaging history updated | 0.85 | 0.563 | 0.571 | 0.10 | - | - |

\*The threshold value for top 80% importance was 1.40.
†The final important features were determined based on the common agreement among the results. Important features were defined as p-values less than 0.05 in either Wald t-test or likelihood ratio test in logistic model, 10 non-zero coefficients in lasso given the proper lambda, and top 80 percentile importance in random forest.
The symbol “-” is used to indicate empty, reference, or 0 coefficients.

**Table 3.** Odds Ratio of Selected Important Features and Pre-selected Clinically Relevant Variables*.

| Variable | OR (95% CI) | *p*-value |
| --- | --- | --- |
| Age (every 1-year increase) | 0.941 (0.927, 0.955) | 0.000 |
| Male (vs. female) | 1.795 (1.129, 2.879) | 0.014 |
| Hypertension | 4.402 (2.469, 8.059) | 0.000 |
| Cerebrovascular disease | 3.612 (1.378, 8.399) | 0.005 |
| Hemoglobin (every 1-g/dL increase) | 0.820 (0.719, 0.936) | 0.003 |
| Positive urine protein | 2.887 (1.861, 4.536) | 0.000 |

\*The reference group for hypertension, cerebrovascular disease, positive urine protein are no history of the condition.

### ESKD and Mortality Outcomes

Among the patients in the RD group, 59 (47.2%) progressed to ESKD with an incidence rate of 91.7 (95% CI, 75.2-111.8) per 1,000 person-years with a medium follow-up time of 4.8 years (Supplemental Table 2). Among patients in the non-RD group, 169 (10.4%) progressed to ESKD with an incidence rate of 17.0 (95% CI, 14.7-19.6) per 1,000 person-years and median follow up time of 6.2 years. The RD group had higher incidence ESKD rates across ages compared to non-RD. After excluding patients >60 years of age, 1,169 patients progressed to ESKD before age 60 with incidence rates 87.2 (95% CI, 70.4-107.9) for RD and 13.3 (95% CI, 10.7-16.6) for non-RD. A total of 19 (15.2%) and 232 (14.3%) patients died in the RD and non-RD groups, respectively. Similar mortality rates were observed between the RD and non-RD (20.5 per 1000-patient years and 21.8 per 1000-person years, respectively).

After excluding those who disenrolled or died within 5-years of incident ADPKD, 976 were identified for analysis of ESKD within 5-years. A total of 31 (38.3%) developed ESKD within 5-years among 81 in the RD group compared with 63 (7.0%) among non-RD (Supplemental Table 3). Cumulative incidence plots for ESKD, ESKD before age 60, and mortality were calculated. RD was highly associated with progression to ESKD during all follow-up years whereas cumulative index plots for mortality were similar between both RD and non-RD (Supplementary Figure 2).

## Discussion

Among patients with ADPKD, there remains a need to identify aggressive phenotypes that can help guide preventative and therapeutic considerations along with resource allocation. Using 42 baseline measurements from routine clinical practice along with longitudinal eGFR data in a large population with ADPKD, we performed multiple data set analyses to identify factors associated with RD and determine a model to further predict patients at risk for RD. Our study found 6 variables that highly predicted RD among patients with ADPKD. We observed younger age of onset, male sex, lower hemoglobin, the presence of proteinuria, hypertension, and cerebrovascular disease to be clinical predictors of RD. We demonstrated 72% sensitivity, 70% specificity, and 70% accuracy in identifying RD.

The European Renal Association-European Dialysis and Transplant Association (ERA-EDTA) algorithm for RD identification amongst ADPKD relies on historical eGFR decline to identify RD. However, using historical eGFR decline alone to identify RD is thought to be of limited value in predicting disease progression in early stages given eGFR decline in ADPKD is nonlinear and often occurs rapidly late in the disease.^14^ In our study, baseline eGFR was higher in the RD group compared to the non-RD group (85.2 vs 72.9 respectively). These findings are consistent with increased hyperfiltration thought to happen first in patients with ADPKD and RD prior to rapid eGFR decline. Baseline creatinine was initially identified as one of the 7 significant variables in identifying RD. However, after including only non-missing data and balancing the clinical relevance and AUC, creatinine and history of liver disease were both excluded in our final modeling tool.

We found age at diagnosis to be significant factors in identifying RD. The CRISP observational study used a large cohort of patients and assessed several prognostic indicators among patients with ADPKD over a six year follow up-period. They similarly found younger age at diagnosis along with total kidney volume (TKV) to be an indicator of early disease progression. Several smaller cohort studies found younger age to be a predictor of RD.^16, 25^ We found presence of proteinuria to also be a significant in identifying RD vs non-RD. This is consistent with prior studies which have shown that urinary protein excretion is correlated with higher mean arterial pressure, larger renal volumes, and increased filtration fraction amongst ADPKD.^25–28^

Total kidney volume (TKV) typically increases continuously from the early stages of the disease and is associated with decline in kidney function. Kidney enlargement progresses significantly in the early stages of the disease making TKV a significant early indicator of disease progression.^16, 29, 30^ One of the most commonly used tools for identifying progression amongst ADPKD is the Mayo Imaging Classification (MIC). The MIC uses height adjusted total kidney volume (htTKV) indexed for age to predict future decline in eGFR. MRIs remain expensive and often difficult to access in many clinical settings.

The PROPKD score uses 4 variables to predict RD: male sex, history of HTN before 35 years of age, first urologic event before 35 years, and PKD mutation type. Similarly, we found male sex and history of hypertension to be 2 of the 6 significant variables in identifying RD in our final modeling tool. PROPKD further relies on identifying PKD mutation type. Genetic factors play an important role in determining severity of ADPKD. PKD1 truncating mutations are associated with the most severe disease with average age of ESKD onset being 56 years whereas PKD1 non-truncating has an average age of ESKD onset of 68 years. PKD2 mutations are associated with the least severe disease with average of ESKD onset being 79 years.^8–13^ Although genetic testing provides prognostic information for ADPKD, it is not often used in routine clinical practice and not widely available on all patients. In addition, ADPKD shows significant phenotypic variability among family members which suggests that disease modifiers including clinical and environmental factors should be considered in evaluating disease progression and prognosis.^8, 31^

The PROPKD score uses both clinical and genetic data to predict likelihood of reaching kidney failure before the age of 60. By assigning points to patients with ADPKD, a score > 6 indicates risk of rapid progression with a 92% chance of reaching kidney failure before age 60. Use of genetic mutation in the scoring system does not take into consideration the intrafamilial variability. A recent long-term follow-up of the CRISP cohort found that while ADPKD genotype was associated with CKD outcomes, it was not considered an independent prognostic factor after adjusting for htTKV.^13^ Similarly, a retrospective study of 164 patients with ADPKD in Spain found the PROPKD score very specific but had low sensitivity in identifying patients with high risk for progression.^15^ Finally, the PROPKD and MIC classification tools were both developed in primarily Caucasian populations. Our clinical risk prediction model is not only an additional tool to help determine RD across a diverse population but also relies on data from routine clinical practice which is often readily available.

Previously, treatment options for ADPKD were limited to management of symptoms and complications.^17^ Management was focused on hypertension, hydration, dietary changes, treatment of pain, urinary tract infections, and ephrolithiasis.^18^ Most patients with ADPKD are diagnosed more than two decades before they reach ESKD.^32, 33^ As novel therapies are developed, there is an increased need for tools that accurately identify higher risk populations which are most likely to benefit from treatment.^34^ While there is no cure for ADPKD, tolvaptan, a selective vasopressin V_2_ receptor blocker, has been FDA approved as the first treatment to slow kidney function in ADPKD. Tolvaptan reduced kidney growth by 45%, reduced eGFR decline by 26% in early ADPKD, and reduced eGFR decline by 35% in advanced ADPKD^19,18^. Newer therapeutic developments further illustrate the need for clinical tools and resources that reliably identify patients eligible for treatment to and guide clinical decision making to help improve outcomes.

## Limitations

There are several potential limitations that may confound the interpretation of our findings. ADPKD was identified using diagnosis codes only which may have misclassified and resulted in over or under-reporting of the ADPKD incidence. However, use of EHRs to identify to rare diseases within KPSC has been described to have modestly high positive predictive values.^35^ Studies evaluating the accuracy of ADPKD diagnosis by ICD codes have demonstrated high sensitivity and positive predictive values exceeding 85%.^36, 37^ In our study, we did not have genetic information nor abdominal imaging with age adjusted TKV total kidney volume for the entire study population. Finally, our study was conducted in a single, integrated healthcare system in one geographic region and may not be generalizable to all patients with ADPKD. Despite these potential limitations, our ADPKD cohort remains one of the largest to date with detailed clinical information capturing laboratory results, medication use, and health care utilizations.

## Conclusion

Among a large, diverse population using real-world data from routine clinical practice, we observed that 6 variables (age, hemoglobin, proteinuria, hypertension, cerebrovascular disease, and sex) highly predict RD among patients with incident ADPKD. Clinical prediction tools may serve as a practical screening tool to capture and manage high-risk patients with ADPKD who may need earlier and more intensive management strategies.

## Data Availability

Data are provided in manuscript and in supplemental attachments. Additional information will be made available upon request to corresponding author.

NA

## Funding

This study was funded by a collaborative grant from Otsuka Pharmaceutical Development & Commercialization, Inc (JJS PI).

## Acknowledgements

The authors would like to thank the KPSC Renal Business Group for their assistance with identifying and providing comprehensive information for the ESKD population.

## Author Contributions

**Study design**: John J. Sim, Yu-Hsiang Shu, KP, Shirin Sundar, Cynthia J. Willey.

**Data acquisition and analysis**: John J. Sim, Qiaoling Chen, Yu-Hsiang Shu, Teresa N. Harrison, Min Young Lee, Mercedes A. Munis, Kerresa Morrissette, KP, Cynthia J. Willey, Shirin Sundar, Ali Nourbakhsh.

**Drafting and submission of the manuscript**: John J. Sim, Simran K. Bhandari, Teresa N. Harrison, Mercedes A. Munis.

**Critical review and revision of manuscript:** Yu-Hsiang Shu, Min Young Lee, Cynthia J. Willey, Shirin Sundar, Kristin Pareja, Kerresa Morrissette, Ali Nourbakhsh.

All authors have given final approval to the manuscript.

## Supplementary Tables

Supplemental Table 1. Mean Lab Values Among Patients in the Non-Rapid Decliner vs. Rapid Decliner Group

Supplemental Table 2. Total Patient Follow-up Time, Events and Incidence Rate of ESKD per 1,000 Person-Years

Supplemental Table 3. ESKD within 5 Years of ADPKD Diagnosis by Non-Rapid Decliner vs. Rapid Decliner Group

## Supplementary Figures

Supplementary Figure 1: Consort diagram

Supplementary Figure 2: Cumulative incidence (CI) plots for ESKD, ESKD < 60 years, and mortality

## Notes

### Competing Interest Statement

Shirin Sundar,Kristin Pareja, and Ali Nourbakhsh are current or former employees of Otsuka Pharmaceutical Development & Commercialization, Inc., Princeton, NJ

### Clinical Trial

NA

### Clinical Protocols

NA

### Funding Statement

Yes

### Author Declarations

The KPSC Institutional Review Board (IRB) reviewed and approved the protocol of this study (#11823).

